# A gain-of-function *GRIA2* variant associated with neurodevelopmental delay and seizures: functional characterization and targeted treatment

**DOI:** 10.1101/2022.03.01.22271646

**Authors:** Ian D. Coombs, Julie Ziobro, Volodymyr Krotov, Taryn-Leigh Surtees, Stuart G. Cull-Candy, Mark Farrant

## Abstract

AMPA-type glutamate receptors (AMPARs) are tetrameric ligand-gated ion channels formed as different combinations of GluA1-4 subunits encoded by the genes *GRIA1-4*. Various pathogenic variants of these genes have been described in patients with developmental delay, intellectual disability, autistic spectrum disorder and seizures. Here we report a heterozygous *de novo* pathogenic missense mutation in *GRIA2* (c.1928 C>T, p.A643V) identified in a one-year-old male patient with seizures, developmental delay and failure to thrive. Electrophysiological investigation of heterologously expressed receptors showed GluA2 A643V to exhibit greatly slowed deactivation, and a markedly reduced extent of desensitization, compared with wild-type GluA2. When GluA2 A643V was coexpressed with the transmembrane AMPAR regulatory protein (TARP) γ2, or with both GluA1 and γ2 to generate TARPed heteromeric receptors, the slowed deactivation and decreased desensitization persisted. We found that the AMPAR negative allosteric modulator perampanel was able to fully block currents from GluA2 A643V/γ2 receptors, albeit with reduced potency compared with wild-type GluA2/γ2. The introduction of perampanel to the patient’s treatment regimen, alongside a modified Atkins diet, was associated with a marked reduction in seizure number, a resolution of failure to thrive, and clear developmental gains. Our study suggests that AMPAR gain-of-function (GoF) underlies the effect of the *GRIA2* variant in our patient, and that perampanel may be beneficial in other patients with *GRIA* GoF variants.

**Author summary:** Rapid communication between brain cells is mediated by the excitatory neurotransmitter glutamate. Mutations affecting genes that encode glutamate-gated ion channels can result in neurodevelopmental disorders and seizures. In a young male patient, we identified a mutation in the gene responsible for production of GluA2, a key subunit of AMPA-type glutamate receptors. We showed that receptors containing GluA2 produced by this *GRIA2* A643V variant were more sensitive to glutamate and gave rise to unusually prolonged or sustained responses, thus causing ‘gain-of-function’. We found that receptors affected by this mutation could, nonetheless, be inhibited by the AMPA receptor-targeting antiseizure medication, perampanel. Prompted by this finding we introduced perampanel alongside the patient’s ongoing antiseizure medication. This coincided with a decrease in the patient’s seizures and clear developmental gains. Taken together, our findings suggest that a *GRIA2* gain-of-function mutation can cause neurological disease, that perampanel can be used to directly counteract the effect of the mutation, and that it may be beneficial in other *GRIA* gain-of-function patients.

## Introduction

AMPA-type glutamate receptors (AMPARs) are ligand-gated cation-permeable ion channels that mediate excitatory synaptic transmission throughout the brain. They are formed as homo- or heterotetrameric assemblies of the subunits GluA1-4, encoded by the genes *GRIA1, GRIA2, GRIA3 and GRIA4* [1]. AMPARs are critical for the correct development of neuronal circuitry and changes in their number or function underlie activity-dependent strengthening or weakening of synaptic signaling and homeostatic adjustments that maintain neuronal excitability [2, 3]. AMPAR-mediated excitatory post-synaptic currents typically exhibit fast activation and decay, allowing them to participate in fast, high-frequency signaling. A rapid deactivation of the synaptic current is generally indicative of low AMPAR affinity and efficient clearance of transmitter from the synaptic cleft, although in situations of persistent activity or slow glutamate clearance, channel closure through desensitization also makes an important contribution. Hence, these two parameters are important measures of AMPAR function.

Given the central importance of AMPARs to excitatory synaptic transmission, it is unsurprising that *GRIA* genes are intolerant to variation. Regions of the transcribed protein that are of particular functional importance are especially intolerant [1], with disease-associated *de novo* variants identified at greatest density in a key conserved motif (SYTANLAAF). This is located towards the extracellular end of the M3 transmembrane helix that forms the extracellular portion of the ion conductance pathway and channel activation gate [4-7]. In general, *GRIA* disorders are typically associated with developmental delay, intellectual disability, autistic spectrum disorder (ASD) and seizures. As yet, however, there have been insufficient patients identified to form clear genotype-phenotype correlations, and there is still a reliance on extended genetic screens or whole genomic analysis to identify cases.

The GluA2 subunit has a central role in determining AMPAR behavior. It is expressed throughout the brain, with GluA2-containing di- or triheteromeric assemblies being the most prevalent AMPAR subtypes [8-11]. Moreover, most GluA2 pre-mRNA undergoes editing at a site that forms the ion selectivity filter of the mature receptor. This editing results in the genetically encoded glutamine (Q) being read as arginine (R) and renders GluA2(R)-containing receptors Ca^2+^-impermeable [12]. *GRIA2* has a Residual Variation Intolerance Score (RVIS) of 10.8% [13] (http://genic-intolerance.org/), indicating that among the healthy population 89.2% of all genes have greater variation. Although *GRIA2* variants in patients with intellectual disability and neurodevelopmental disorders have been reported to produce predominantly AMPAR loss-of-function [7], gain-of-function variants have nonetheless been identified in other GluA subunits [5, 6]. Accordingly, electrophysiological investigation is required to assess the functional effects of novel variants. The information from such studies may enable a precision medicine approach to treatment. In this regard, while no AMPAR positive modulators are currently available in the clinic, perampanel, a selective AMPAR negative allosteric modulator (NAM) and licensed anti-epileptic drug [14], could be beneficial for *GRIA* patients with identified gain-of-function variants.

We describe a patient with a novel *GRIA2* variant in the SYTANLAAF motif who presented with symptoms including seizures, developmental delay and failure to thrive. Seizures were not eliminated by phenobarbital and levetiracetam nor by trials of additional valproic acid or clobazam, and a modified Atkins diet was subsequently introduced. Here we establish that the variant produces a gain-of-function, conferring slowed AMPAR deactivation, decreased desensitization and increased glutamate potency. Accordingly, we examined the blocking effect of perampanel. Although the drug displayed reduced potency against variant receptors, it was nonetheless a fully effective inhibitor at high doses. Following the addition of perampanel to the patient’s treatment plan he demonstrated improved seizure control, resolution of failure to thrive, and has made clear developmental gains.

## Results

### Clinical findings in a patient with *GRIA2* A643V

The male patient in this study was born at 35-weeks’ gestation, following a pregnancy complicated by polyhydramnios. Neurological evaluation was performed at 7 months of age following episodes of back arching and vomiting after feeds. His prior history was notable for gastro-esophageal reflux disease; there was no relevant family medical history. His exam showed decreased muscle tone and mild developmental delay. The EEG at that stage appeared normal (**Fig 1a**).

**Figure 1.**
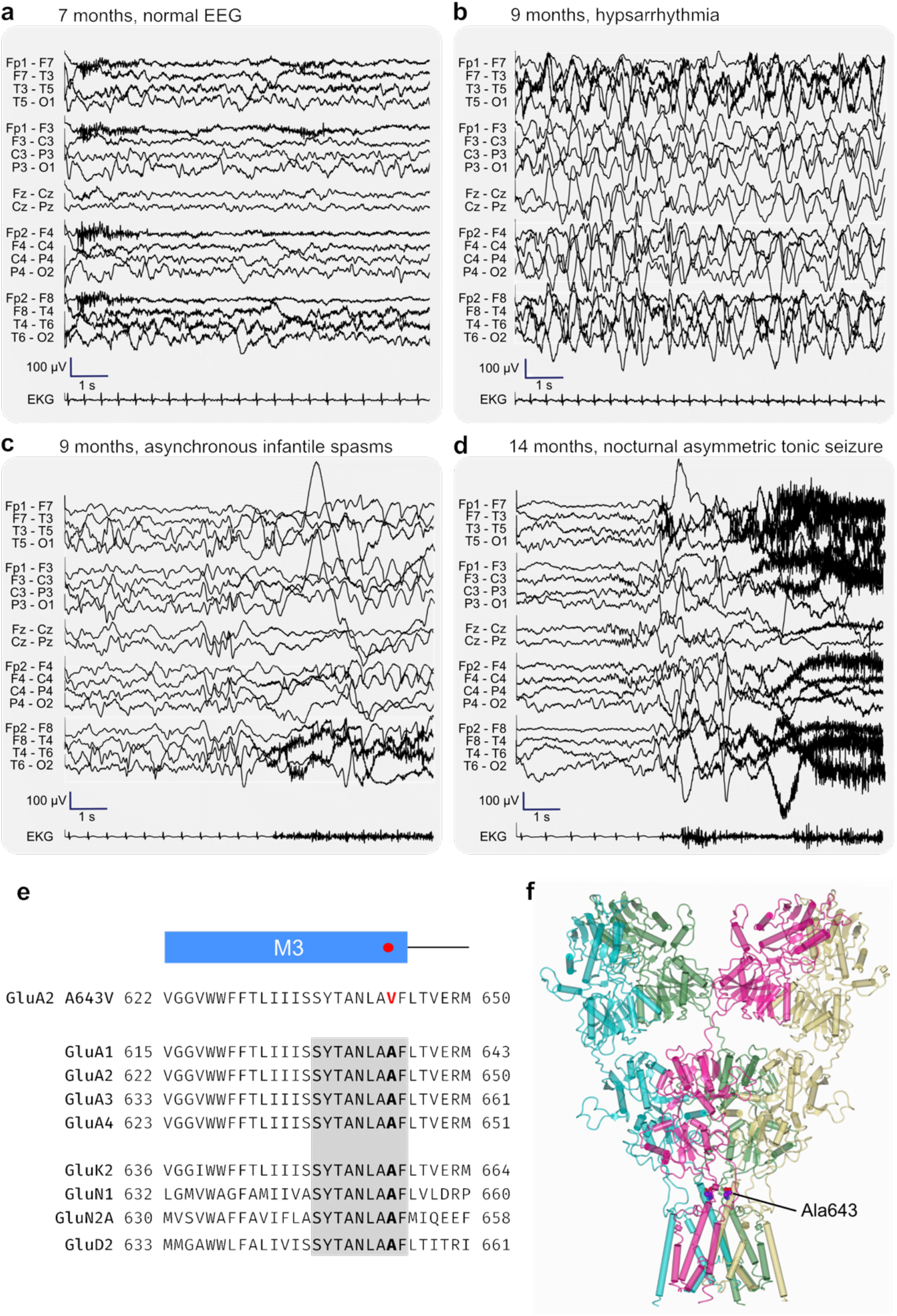
Clinical presentation of a patient with the variant *GRIA2* A643V. **a-d**) EEG findings. Progression from normal EEG at 7 months of age (a) to hypsarrhythmia (b) with asynchronous infantile spasms (c) at 9 months of age. Hypsarrhythmia resolved with ACTH therapy, but frequent nocturnal asymmetric tonic seizures (d) developed at 14 months of age. **e**) Sequence alignments highlighting the position of Ala643 in the third membrane region (M3) of the AMPAR subunit. The surrounding region is completely conserved between all four AMPAR subunits, and the SYTANLAAF motif (gray box) is conserved throughout the iGluR superfamily. The gene sequences are from Human GluA1 (NP_001107655.1), Human GluA2 (NP_000817.5), Human GluA3 (NP_015564.5), Human GluA4 (NP_000820.4), Human GluK2 (NP_068775.1), Human GluN1 (NP_015566.1), Human GluN2A (NP_000824.1), and Human GluD2 (NP_001501.2). **f**) Three-dimensional structure of a GluA2 homomeric receptor (PDB ID: 5L1F; [16]) with the position of Ala643 indicated (shown as spheres on all four subunits).

At 9 months of age the patient presented to the emergency department with a fever, decreased interaction and new symptoms consisting of dystonic posturing with back arching and upward eye deviation followed by generalized body limpness. Continuous EEG monitoring provided a diagnosis of atypical infantile spasms with myoclonic seizures and a hypsarrhythmia background, with a BASED (Burden of Amplitudes and Epileptiform Discharges) score of 4 [15] (**Fig 1b,c**). Infantile spasms and hypsarrhythmia were resolved by treatment with ACTH (adrenocorticotropic hormone) and levetiracetam. Brain imaging was performed, with the head CT showing notable extra-axial hyperdensity along the anterior falx. Potentially, this was indicative of a subdural hematoma, which was confirmed by brain MRI and vascular imaging (**Fig S1**). Further, the MRI exhibited prominent bi-frontal extra-axial space and restricted diffusion along the left insular cortex and left medial temporal lobe (**Fig S1**). We found no evidence for infectious or traumatic etiologies. At this point, Glutaric aciduria type 1 (GA1) was considered and carnitine supplementation was initiated. We performed an extended epilepsy gene panel, to assess for GA1 (the *GCDH* gene), in addition to other causes of early infantile epileptic encephalopathies (see below).

The patient was subsequently admitted at 11 months of age for sub-clinical focal status epilepticus in the setting of respiratory syncytial virus (RSV) infection, requiring introduction of phenobarbital. His global developmental delays became more pronounced during this time period, and asymmetric tonic seizures from sleep developed at 14 months of age (**Fig 1d**). Seizures occurred every 7-10 days and generally clustered (up to 12 nightly seizures). Meanwhile, he developed significant vomiting and constipation, resulting in failure to thrive – his weight fell from the 63%-ile at 6 months of age to the 0.07%-ile by 20 months of age. Seizures were not eliminated by trials of additional valproic acid or clobazam, and a modified Atkins diet was subsequently introduced. This diet was later supplemented with medium-chain triglycerides (MCTs).

Our genetic analysis revealed a heterozygous variant in *GRIA2* (c.1928 C>T, p.A643V, NM_001083619.1) with no other variants of clinical interest (**Fig 1e**). The variant was novel, being absent from the Genome Aggregation Database (gnomAD) and the ClinVar database and was confirmed as *de novo* by trio sequencing. *GRIA2* Ala643 is located at the *lurcher* site [17] in the highly conserved sequence (SYTANLA<u>A</u>F) that forms the upper AMPAR ion channel gate (**Fig 1f**). Bioinformatic predictions suggested that the variant was likely to be pathogenic – SIFT pathogenic, score 0 [18]; polyphen 2, probably damaging, score 0.999 [19]; VARITY R, score 0.953 [20]. Indeed, variants at this site have previously been suggested as disease-causing for *GRIA1* [6], *GRIA4* [4], and *GRIA3* (present in ClinVar database) (**Fig 1f**). Previous examination of *GRIA2* missense variants underlying neurodevelopmental disorders and seizures, found them to cause primarily a loss-of-function (LoF) [7]. However, mutation at the analogous site in *GRIA1* (A636T) produces a gain-of-function (GoF) due to destabilization of the closed gate [17] (**Fig 1f**). It was therefore crucial to understand the impact of the valine variant on AMPAR function.

### GluA2 A643V causes gain-of-function

To determine the functional effect of the *GRIA2* A643V variant we expressed GluA2(Q) wild-type or A643V subunits in HEK293 cells and compared their responses to rapid glutamate application in outside-out patches (**Fig 2a-c)**. The homomeric GluA2(Q) A643V receptors were functional and produced currents with distinct kinetic features. For currents evoked using short glutamate applications (10 mM, –60 mV, 1-2 ms) the weighted time constant of deactivation (τ_w, deact_) was roughly 7-fold greater with A643V compared to wild-type (**Table 1**) (**Fig 2a,d**). With long glutamate applications (10 mM, –60 mV, 500 ms) we found that the A643V variant markedly affected AMPAR desensitization. The weighted time constant (τ_w, des_) was almost double that of wild-type and the fractional steady-state current (*I*_ss_) was increased more than 50-fold, indicating that the extent of desensitization was greatly reduced (**Table 1**) (**Fig 2b,e**). Consistent with this large steady-state current, the recovery from desensitization of the variant was more than twice as fast as that of the wild-type (**Table 1**) (**Fig 2c,f**). Taken together, these results indicate that A643V is a GoF variant. We found no evidence that the variant altered GluA2 pore properties, as non-stationary fluctuation analysis revealed no effect on weighted mean channel conductance or peak open probability (**Table 1**) (**Fig 2b,g**).

**Figure 2.**
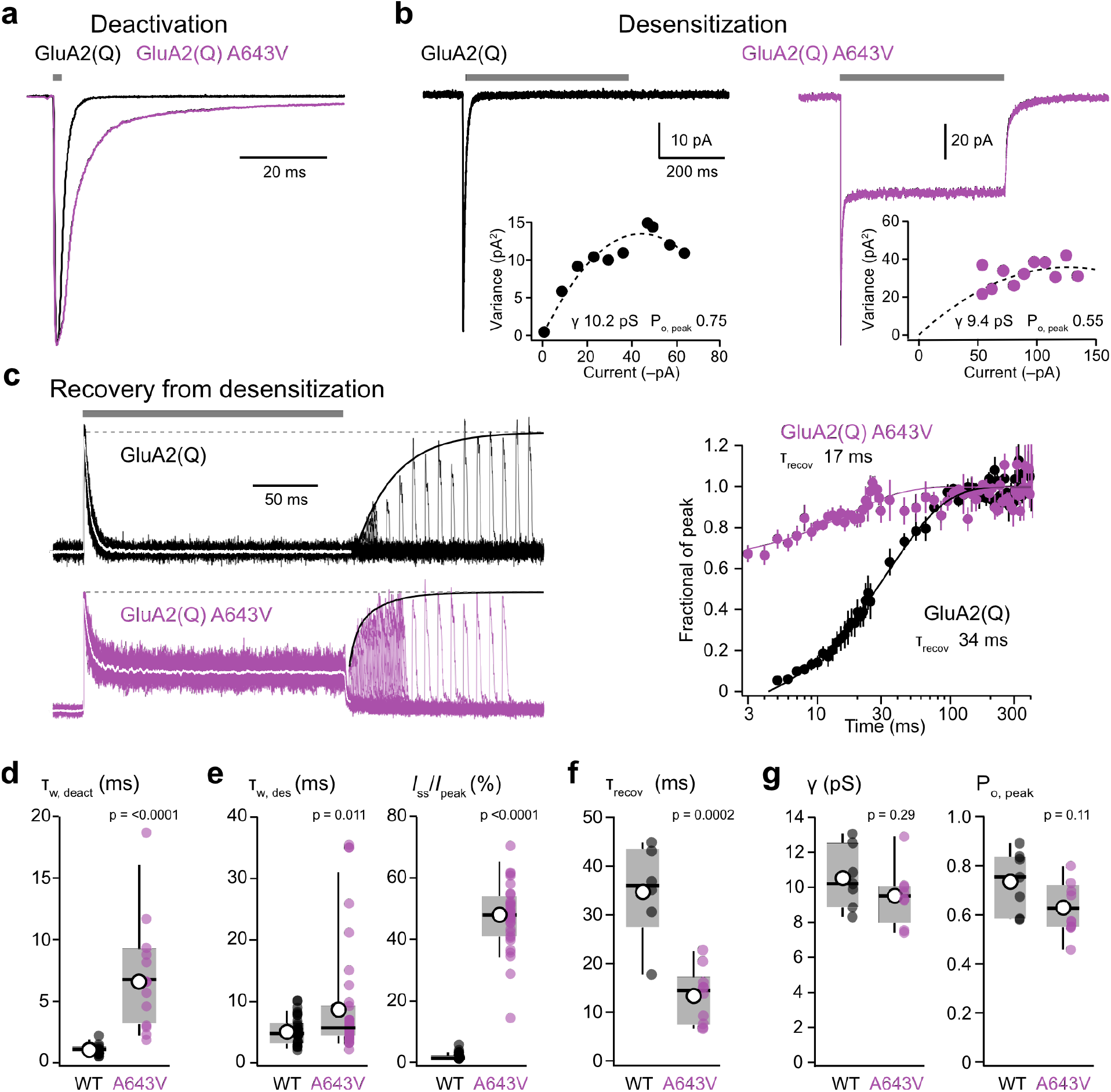
The *GRIA2* A643V variant generates a gain-of-function. **a**) Representative outside-out patch responses (10 mM glutamate, 1 ms, −60 mV; gray bar) from HEK293 cells transfected with wild-type (WT) GluA2(Q) (black) or GluA2(Q) A643V (purple) (superimposed). **b**) Representative outside-out patch responses to 500 ms applications of glutamate (10 mM, −60 mV). Insets show corresponding current-variance relationships and estimated channel conductance (γ) and peak open probability (*P*_o, peak_). **c**) Representative outside-out patch responses showing recovery from desensitization; superimposed currents (inverted for display) evoked by steady-state glutamate application (200 ms, −60 mV; gray bar) and subsequent 10 ms applications of glutamate, applied at increasing intervals. Averaged currents before recovery shown as white traces. Solid black lines are exponential fits of peak current recovery. Right; pooled recovery timecourse plot for GluA2(Q) (n = 10) and GluA2(Q) A643V (n = 6). Symbols and error bars indicate mean values with s.e.m., and the solid lines are exponential fits, yielding the indicated τ_recov_ values. **d**) Pooled τ_w, deact_ data for GluA2(Q) (n = 16) and GluA2(Q) A643V (n = 13). Box-and-whisker plots indicate the median (black line), the 25–75th percentiles (box), and the 10–90th percentiles (whiskers); filled circles are data from individual patches and open circles indicate means. **e**) Pooled τ_w, des_ and *I*_ss_ data for GluA2(Q) (n = 31) and GluA2(Q) A643V (n = 33) from 500 ms glutamate applications. **f**) Pooled τ _recov_ data for GluA2(Q) (n = 6) and GluA2(Q) A643V (n = 10). **g**) Pooled weighted mean conductance (γ) and *P*_o, peak_ data for GluA2(Q) (n = 8) and GluA2(Q) A643V (n = 7). Indicated p-values are from two-sided approximate permutation t-tests comparing wild-type (WT) and A643V variant (**Table 1**).

**Table 1.** Summary of GluA2 A643V effects on homomeric and heteromeric AMPARs. Data are presented as mean ± standard error (s.e.m.) from n patches. Unpaired mean differences are given with their 95% bias corrected and accelerated (BCa) confidence intervals [upper bound, lower bound], calculated from 5000 bootstrap resamples. All p-values were calculated using a non-parametric two-sided approximate permutation t-test, with 10000 bootstrap replicates. The p-values are reported as equalities, unless < 0.0001.

|  | GluA2(Q) | GluA2(Q) A643V | Unpaired mean difference [95%CI] | p-value |
| --- | --- | --- | --- | --- |
| $T_{w, deact}$ (ms) | 1.0 ± 0.1 (16) | 6.9 ± 1.3 (13) | 5.9 [4.0, 9.0] | < 0.0001 |
| $T_{w, des}$ (ms) | 5.1 ± 0.4 (31) | 9.5 ± 1.7 (33) | 4.4 [1.7, 8.3] | 0.011 |
| $I_{ss}/I_{peak}$ (%) | 0.9 ± 0.2 (31) | 47.4 ± 2.0 (33) | 46.5 [42.2, 50.3] | < 0.0001 |
| $T_{recov}$ (ms) | 34.7 ± 4.0 (6) | 13.4 ± 1.8 (10) | -21.3 [-28.0, -12.1] | 0.0002 |
| Conductance (pS) | 10.5 ± 0.7 (7) | 9.5 ± 0.6 (8) | -1.0 [-2.6, 0.7] | 0.29 |
| $P_{o, peak}$ | 0.74 ± 0.12 (7) | 0.63 ± 0.11 (8) | -0.11 [-0.21, 0.01] | 0.11 |
| Glu $EC_{50, ctz}$ (μM) | 322 ± 63 (5) | 59 ± 9 (6) | -263 [-402, -177] | < 0.0001 |
| Glu $EC_{50, peak}$ (mM) | 1.14 ± 0.20 (5) | 0.15 ± 0.02 (5) | -0.99 [-1.29, -0.59] | < 0.0001 |
|  | GluA1/2(R) | GluA1/2(R) A643V |  |  |
| $T_{w, deact}$ (ms) | 2.0 ± 0.4 (7) | 18.6 ± 6.8 (6) | 16.6 [7.8, 36.8] | 0.0007 |
| $T_{w, des}$ (ms) | 7.2 ± 1.0 (9) | 32.7 ± 8.5 (5) | 25.5 [15.3, 50.4] | 0.0002 |
| $I_{ss}/I_{peak}$ (%) | 2.6 ± 1.0 (9) | 34.2 ± 8.7 (5) | 31.6 [15.1, 45.6] | 0.0005 |

We next examined the effect of A643V on glutamate potency. We first recorded responses to various glutamate concentrations in the presence of the positive allosteric modulator (PAM) cyclothiazide to limit desensitization (**Fig 3a, b**). We found that glutamate displayed increased potency at A643V AMPARs, with the concentration producing half-maximal current (*EC*_50, CTZ_) being reduced by 5-fold (**Table 1**). We also examined the concentration dependence of glutamate-evoked peak currents in the absence of cyclothiazide (**Fig 3c, d**). where again the variant increased glutamate potency (**Table 1**). These results are consistent with A643V being a GoF variant.

**Figure 3.**
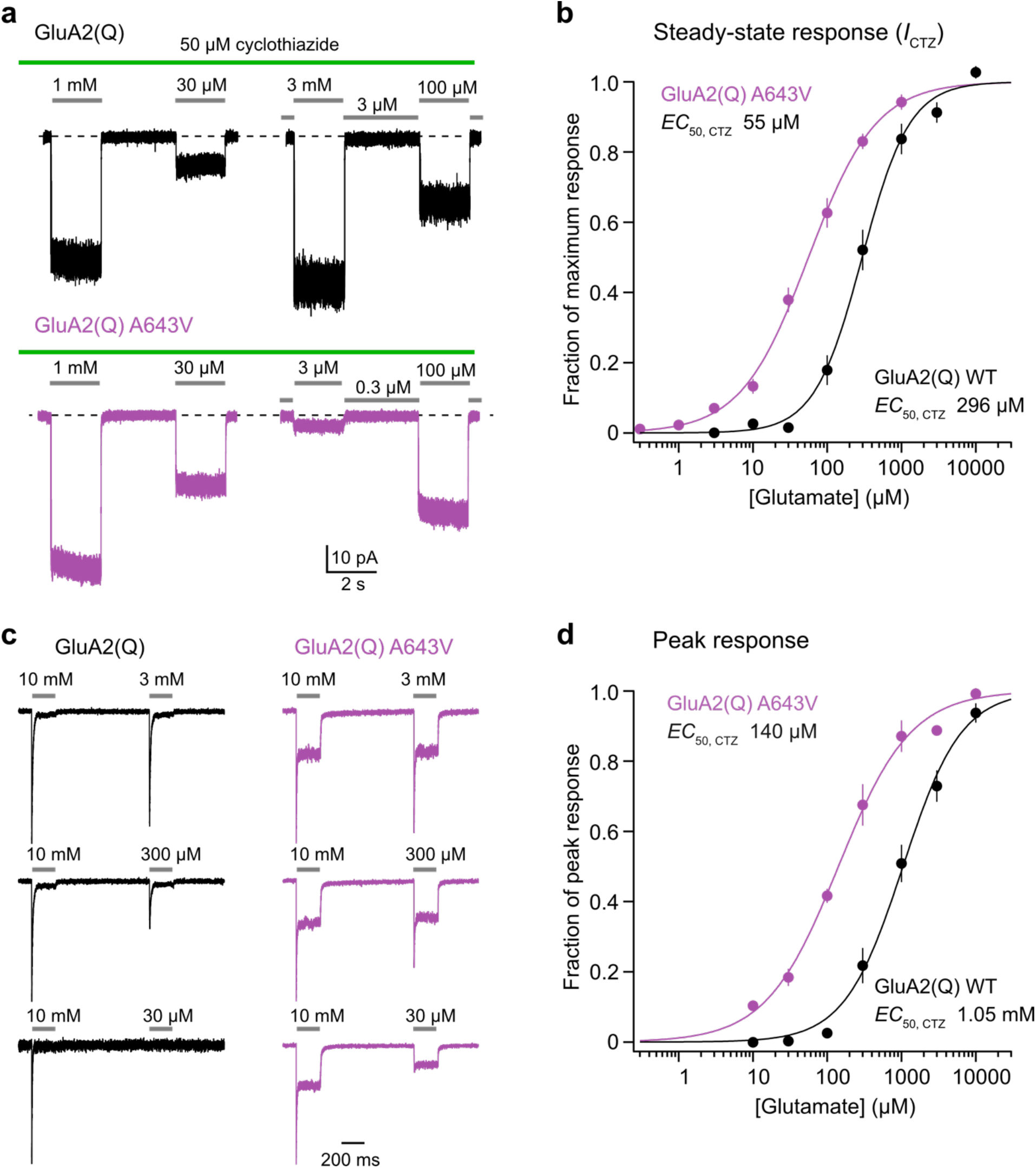
The *GRIA2* A643V variant increases glutamate potency. **a**) Representative currents evoked by different concentrations of glutamate (gray bars) applied to outside-out patches from HEK293 cells transfected with wild-type (WT) GluA2(Q) (black) or GluA2(Q) A643V (purple) in the presence of 50 μM cyclothiazide (green bar). **b**) Pooled normalized concentration-response curves for glutamate in the presence of cyclothiazide for GluA2(Q) (n = 5) and GluA2(Q) A643V (n = 6). Symbols and error bars indicate mean values with s.e.m., and the solid lines are fits to the Hill equation, yielding the indicated *EC*_50_ values. **Table 1** reports the statistical analysis of *EC*_50_ values obtained from separate fits of the data from individual patches. **c**) Representative glutamate responses in the absence of cyclothiazide. Each sweep contained an initial jump into 10 mM glutamate, for normalization, followed by a jump into a lower glutamate concentration. **d**) Pooled normalized peak-current concentration-response curves for GluA2(Q) (n = 5) and GluA2(Q) A643V (n = 5) (details as in b).

Having established that GluA2 A643V produces a GoF in homomeric Q/R unedited receptors, we next examined its impact when incorporated into heteromeric receptors. Specifically, we co-expressed GluA1 and Q/R edited GluA2, the AMPAR subunits that predominate in the hippocampus [10, 21]. As these preferentially co-assemble into tetramers containing two copies of GluA1 and two copies of GluA2, we speculated that the functional effect of the A643V variant might be weakened in the presence of GluA1. However, when we recorded currents at +60 mV with 100 μM NASPM in the intracellular (pipette) solution to block any GluA1 homomers [22], we found that the variant slowed deactivation and markedly reduced desensitization (**Fig 4a, b**). Thus, τ_w, deact_ was increased nearly 10-fold, τ_w, des_ was increased more than 4-fold, and *I*_ss_ was increased by more than 10-fold (**Table 1**). Taken together, these data show that GluA2 A643V imparts a marked GoF when the variant is part of a heteromeric GluA1/2 assembly.

**Figure 4.**
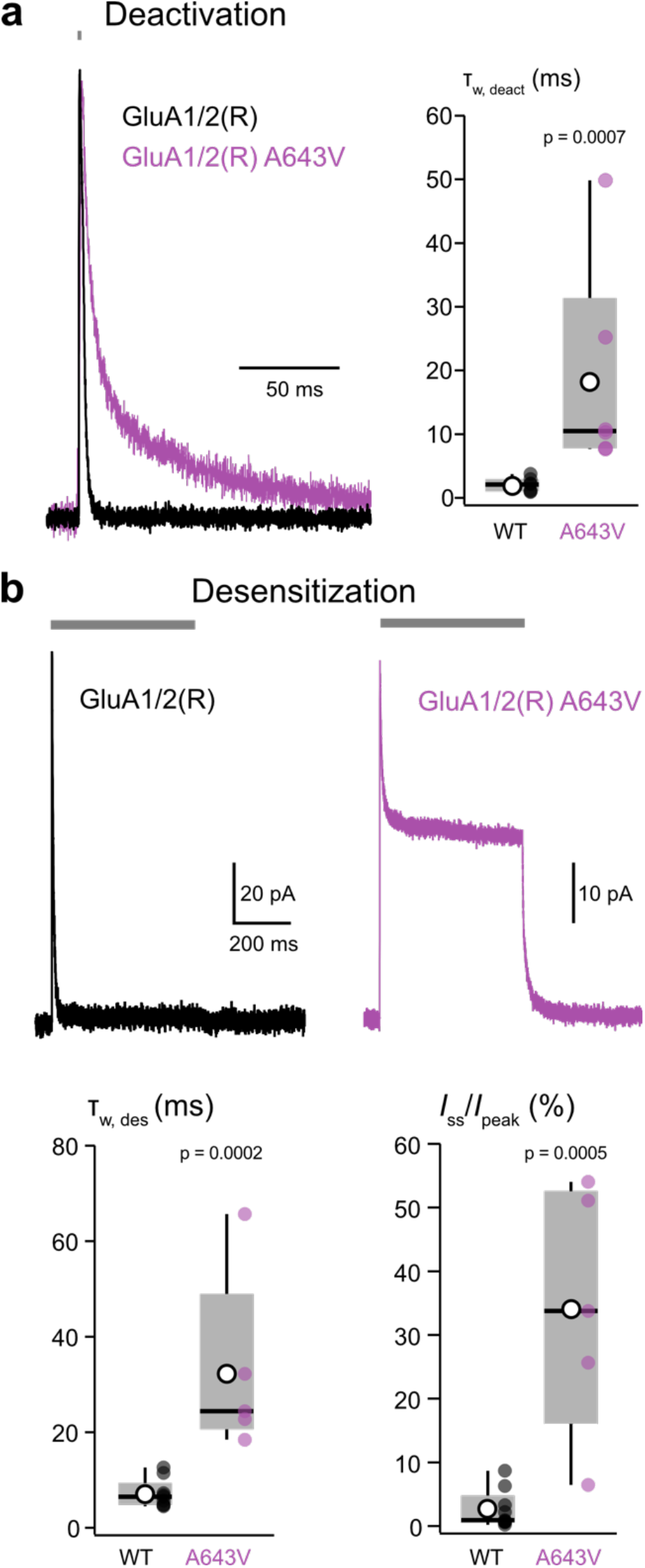
*GRIA2* A643V alters the properties of GluA1/2 heteromeric AMPARs. **a**) Representative deactivating outside-out patch responses (10 mM glutamate, 1 ms, +60 mV; gray bar) from HEK293 cells transfected with GluA1 and GluA2(R) (black) or GluA1 and GluA2(R) A643V (purple) (superimposed). Right: pooled τ_w, deact_ data for wild-type (WT) GluA1/GluA2(R) (n = 7) and GluA1/GluA2(R) A643V (n = 6). Boxplots as in Fig 2. **b**) Representative desensitizing responses (10 mM glutamate, 500 ms; gray bar) from WT and A643V heteromers. **c**) Pooled τ_w, des_ and *I*_ss_ data for GluA1/GluA2(R) (n = 9) and GluA1/GluA2(R) A643V (n = 5). Indicated p-values are from two-sided approximate permutation t-tests comparing wild-type (WT) and A643V variant (**Table 1**).

Native AMPAR assemblies contain auxiliary proteins, the most prevalent being the transmembrane AMPAR regulatory proteins (TARPs) [23, 24]. As these affect many biophysical and pharmacological AMPAR properties [25], we next considered the effect of the A643V variant on AMPAR assemblies that contained the prototypical TARP γ2 (stargazin). Responses to short glutamate applications showed that, as seen in absence of TARP, τ_w, deact_ was increased for both homomeric and heteromeric receptors (∼10- and 3-fold, respectively) (**Fig 5a,b**) (**Table 2**). In response to long glutamate applications the desensitization of GluA2(Q)/γ2 and GluA1/2(R)/γ2 was not slowed by the presence of the variant. However, *I*_ss_ was greatly increased (**Fig 5c,d**) (**Table 2**). Thus, in terms of slowed deactivation and decreased steady-state desensitization, GluA2 A643V produces GoF in all the AMPAR assemblies we examined.

**Figure 5.**
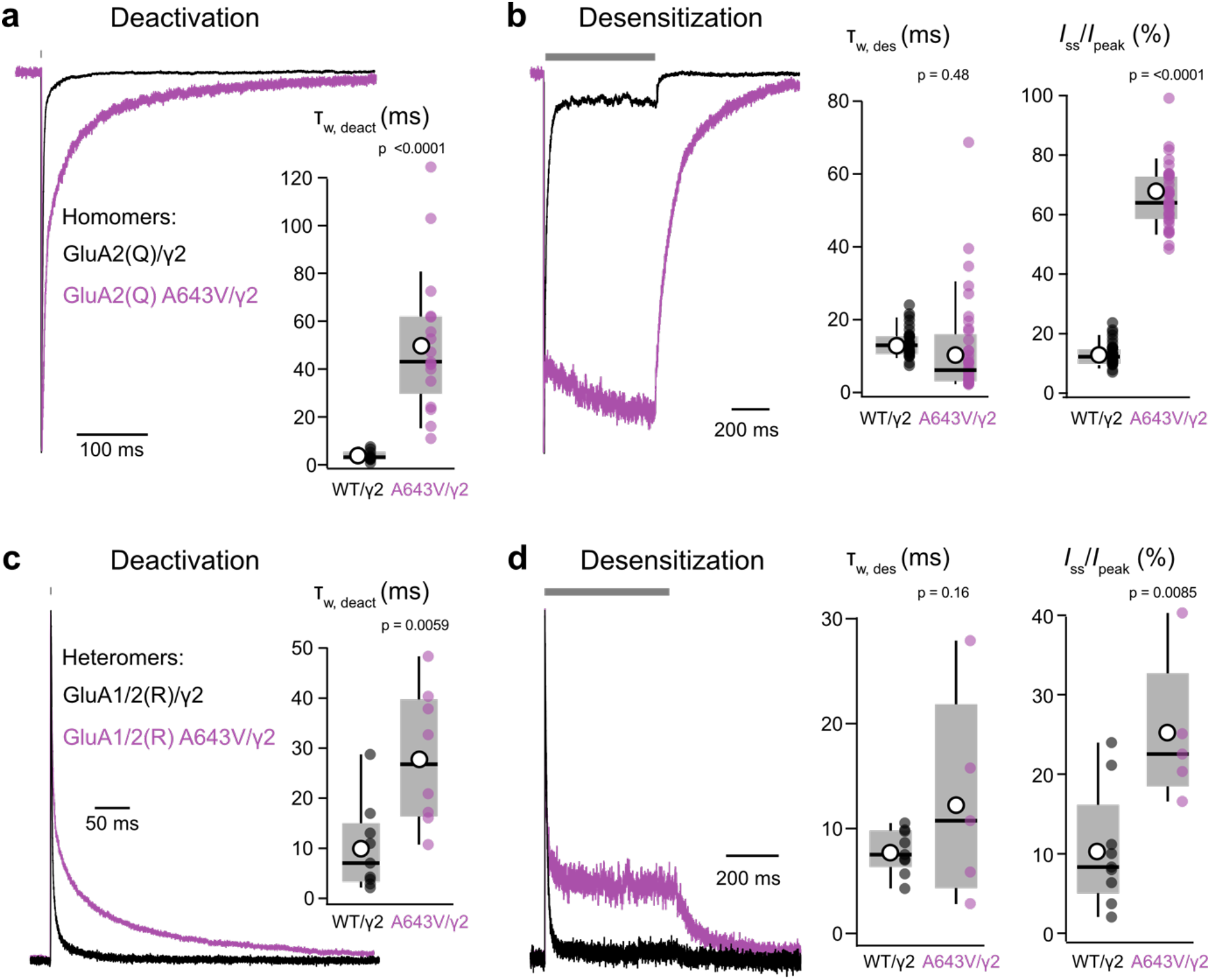
*GRIA2* A643V alters the properties of TARP-associated homomeric and heteromeric AMPARs. **a**) Representative superimposed deactivating outside-out patch responses (10 mM glutamate, 1 ms, −60 mV; gray bar) from HEK293 cells transfected with GluA2(Q) and TARP γ2 (black) or GluA2(Q) A643V and γ2 (purple). Inset: pooled τ_w, deact_ data for wild-type (WT)/γ2 and A643V/γ2 (box-and-whisker plot as in Fig 2). **b**) Representative superimposed desensitizing outside-out patch responses (10 mM glutamate, 500 ms, −60 mV; gray bar) from GluA2(Q)/γ2 (black) and GluA2(Q) A643V/γ2 (purple). Right: pooled τ_w, des_ and *I*_ss_ data **c**) Representative superimposed deactivating currents from heteromeric receptors with TARP γ2; GluA1/GluA2(R)/γ2 (black) or GluA1/GluA2(R) A643V/γ2 (purple) (10 mM glutamate, 1 ms, +60 mV; gray bar). Inset: pooled τ_w, deact_ data. **d**) Representative superimposed desensitizing responses (10 mM glutamate, 500 ms, +60 mV) from GluA1/GluA2(R)/γ2 (WT; black) and GluA1/GluA2(R) A643V/γ2 (purple). Right: pooled τ_w, des_ and *I*_ss_ data. Indicated p-values are from two-sided approximate permutation t-tests comparing wild-type (WT) and A643V variant (**Table 1**).

**Table 2.** Summary of GluA2 A643V effects on homomeric and heteromeric AMPARs expressed with TARP γ2. Data are presented as mean ± standard error (s.e.m.) from n patches. For GluA2(Q) A643V/γ2, the perampanel (PER) *IC*_50_ is shown for both the initial peak and steady-state (s-s) currents. Details as in Table 1.

| | GluA2(Q)/γ2 | GluA2(Q) A643V/γ2 | Unpaired mean difference [95%CI] | p-value |
| --- | --- | --- | --- | --- |
| $T_{w, deact}$ (ms) | 3.6 ± 0.6 (11) | 50.5 ± 7.2 (17) | 46.9 [35.9, 64.1] | < 0.0001 |
| $T_{w, des}$ (ms) | 13.6 ± 0.8 (29) | 11.6 ± 2.3 (36) | -2.0 [-5.6, 4.2] | 0.48 |
| $I_{ss}/I_{peak}$ (%) | 14.1 ± 2.2 (29) | 66.2 ± 1.7 (37) | 52.1 [46.2, 57.2] | < 0.0001 |
| PER $IC_{50 peak}$ (μM) | 0.13 ± 0.02 (6) | 1.29 ± 0.38 (4) | 1.17 [0.66, 1.98] | 0.0052 |
| PER $IC_{50 s-s}$ (μM) | | 3.08 ± 0.26 (4) | | |
| | GluA1/2(R)/γ2 | GluA1/2(R) A643V/γ2 | | |
| $T_{w, deact}$ (ms) | 10.0 ± 2.9 (9) | 28.0 ± 4.8 (8) | 18.0 [7.71, 28.2] | 0.0059 |
| $T_{w, des}$ (ms) | 7.8 ± 0.7 (9) | 12.6 ± 4.4 (5) | 4.8 [-1.53, 14.7] | 0.16 |
| $I_{ss}/I_{peak}$ (%) | 10.5 ± 2.5 (9) | 25.0 ± 4.1 (5) | 14.4 [6.9, 23.9] | 0.0085 |

### Perampanel inhibition of GluA2 A643V

As we found the A643V mutation to cause a clear GoF, we next sought to examine the effect of the AMPAR NAM perampanel, a licensed anti-epileptic drug [14]. For both GluA2(Q)/γ2 and GluA2(Q) A643V/γ2 perampanel inhibited the response to glutamate (**Fig 6a**). However, perampanel was roughly 10 times less potent against the variant than the wild-type (when assessing its effects on the initial peaks of the glutamate-evoked currents) (**Fig 6b**) (**Table 2**). In some recordings the response of GluA2(Q) A643V/γ2 in the presence of perampanel showed a slow ‘run-up’ (**Fig 6b**). When assessing the steady-state responses, rather than the initial peaks, the perampanel potency was reduced (**Table 2**). Of note the perampanel-bound crystal structure of GluA2(Q) [16] shows that the perampanel binding site is adjacent to the channel gate and the Ala643 residue, potentially accounting for the decreased potency of the drug against the variant receptors (**Fig 6c**).

**Figure 6.**
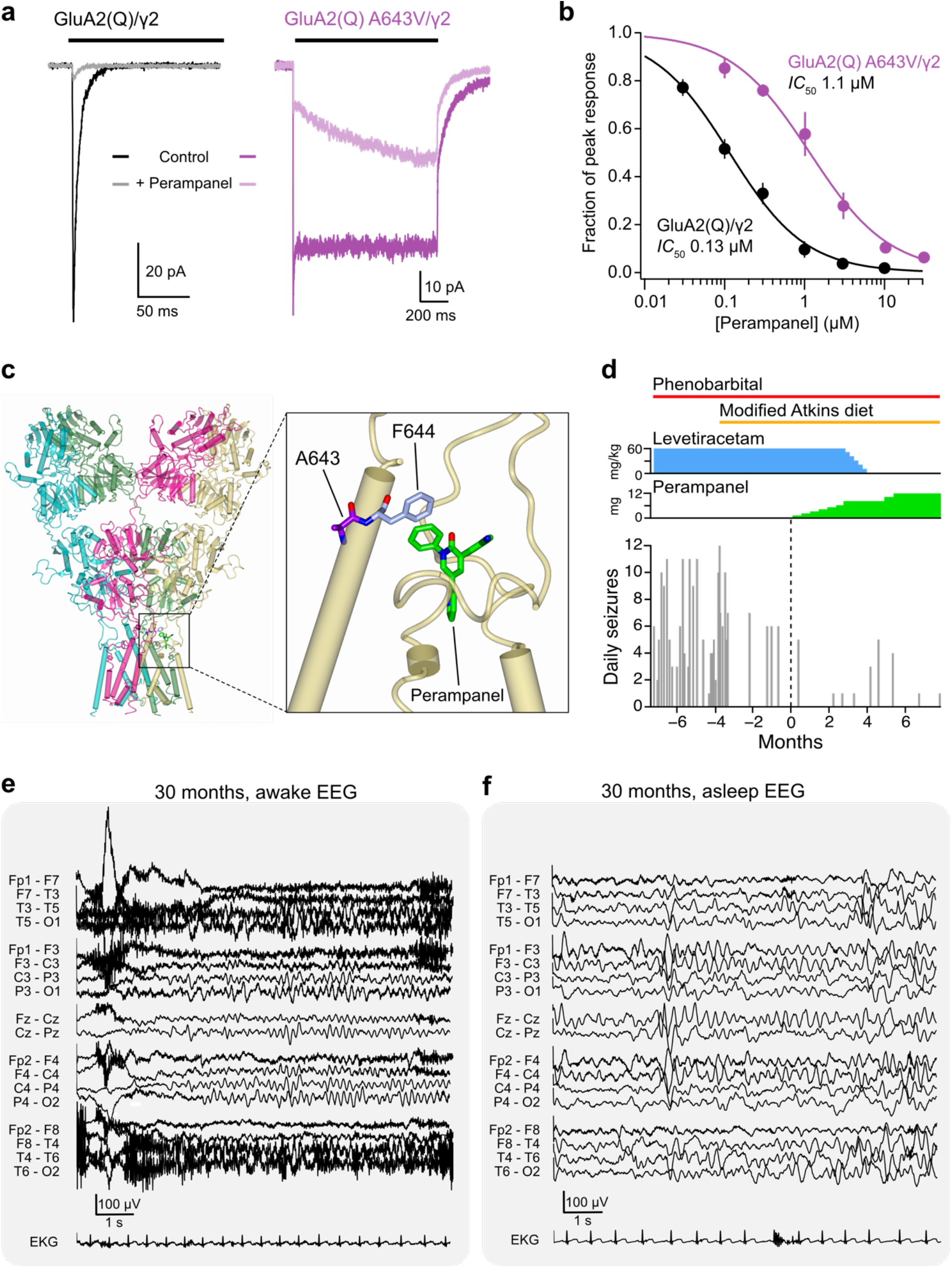
Functional and clinical impact of perampanel. **a**) Representative GluA2(Q)/γ2 (black) and GluA2(Q) A643V/γ2 currents (purple) with inhibition by 3 μM perampanel (pale traces). Note the run-up in steady-state current seen with the variant. **b**) Concentration inhibition curves demonstrating decreased potency of PMP against the A643V variant. Symbols and error bars indicate mean values with s.e.m., and the solid lines are fits to the Hill equation, yielding the indicated *IC*_50_ values. **Table 2** reports the statistical analysis of *IC*_50_ values obtained from separate fits of the data from individual patches. **c**) Crystal structure of a GluA2(Q) receptor with perampanel bound, showing the binding site adjacent to A643, and the F644 that interacts the phenyl ring of perampanel (PDB: 5L1F [16]). **d**) Number of parent-reported daily seizures in the months before and after commencement of perampanel treatment (at 0 months). Colored bars and plots denote the timing of anti-seizure medications. Phenobarbital (6 mg/kg daily) is currently being weaned, while the Modified Atkins diet is being maintained. Following the introduction of perampanel, levetiracetam was gradually withdrawn. **e, f**) EEG at 30 months of age following treatment with perampanel for 3 months. The awake (e) and sleep (f) interictal patterns remain slow and disorganized, with multifocal spikes in sleep, consistent with an epileptic encephalopathy with ongoing risk for seizures.

### Clinical introduction of perampanel

Following the introduction of the modified Atkins diet at 24 months the patient displayed an initial drop in seizure number and frequency, although occasional clusters of 6 or more seizures persisted (**Fig 6d**). Our functional data demonstrate that, while perampanel has a reduced potency against the variant, it is an effective blocker at high doses. Following confirmation of a *GRIA2* GoF mechanism, adjunctive treatment with perampanel was initiated at 27 months of age. Perampanel was introduced at 1 mg daily, with gradual up-titration of the dose. Perampanel introduction was associated with a further reduction in seizure frequency, allowing the patient to be successfully weaned from levetiracetam and he is gradually being weaned from phenobarbital. At 32 months of age the modified Atkins was supplemented with MCT oil [26].

Following the introduction of perampanel the rate of developmental gains, as reported by the patient’s occupational therapist, accelerated. He gained the ability to sit independently and weight-bear in quadruped and is ambulating with a gait-trainer. Fine motor progression was also noted, including gains in grasping and reaching. His visual attention has significantly improved in terms of both eye contact and facial recognition. He is developing communication skills, and can both follow simple commands and verbally respond consistently to “yes” and “no” questions. Constipation, vomiting and failure to thrive have resolved and current weight (34 months) has recovered (from the 0.07%-ile at 20 months) to the 15.8%-ile. The patient has suffered no adverse effects of perampanel and has been successfully titrated to the maximum dose of 12 mg daily. His family reports a significant improvement in baseline irritability, and in particular, improved mood with consistent smiling and purposeful laughing following his perampanel dose, behaviors that were not present prior to perampanel. Nonetheless, the EEG performed at 30 months (3 months after starting perampanel, and 6 months after the onset of the modified Atkins diet) remained slow and disorganized, with multifocal spikes predominantly in the bilateral posterior and left temporal regions, suggestive of an epileptic encephalopathy with ongoing risk for recurrent seizures (**Fig 6e,f**). Consistent with this, two additional clusters of asymmetric tonic seizures associated with a viral illness and teething, respectively, were observed 4-5 months after perampanel initiation. These were successfully rescued with additional phenobarbital.

## Discussion

### *GRIA* disorder

*GRIA* disorder is an emerging neurological disease with over 100 published variants identified across the four *GRIA* genes [1]. Most patients with *GRIA* disorder display intellectual disability and developmental delay, accompanied, in many cases, with ASD and seizures. However, beyond these broad groupings there is great variety in the nature and onset of symptoms. Our patient exhibited some of the features previously reported for various different individuals with *GRIA2* disorder: for example, in the onset of symptoms at ∼6 months, the presence of myoclonic seizures and developmental delay [7]. However, certain of our patient’s symptoms had not previously been reported; these include failure-to-thrive, associated with gastrointestinal symptoms. Our study establishes that *GRIA2* A643V is a GoF variant – a previously unreported molecular phenotype which could be expected to account for the neurological symptoms observed.

The diverse symptoms of *GRIA* disorder likely stem from the identity of the AMPAR subunit involved in each case and the varied functional impact of the various mutations. Where functional analysis of variants has been performed, studies have identified both GoF changes (in *GRIA1* [6] and *GRIA3* [27, 28], and LoF changes (in *GRIA2* [7] and *GRIA3* [5, 29, 30]. For *GRIA2*, a wide variety of causative defects have been described. These include missense [7, 31], nonsense [7, 32], frame shift, truncation, splice site and in-frame deletion variants [7], as well as haploinsufficiency [7, 33-35]. Of the ten *GRIA2* missense variants that have been functionally examined, six were reported to be LoF, three displayed behavior indistinguishable from that of wild-type, while one (a variant at the Q/R site) led to the production of Ca^2+^-permeable GluA2-containing receptors [7]. Our case represents the first identification of a *GRIA2* GoF variant featuring prolonged activation and increased glutamate potency.

The A643V variant affects the most highly conserved sequence in the ionotropic glutamate receptor superfamily, namely the SYTANLA<u>A</u>F motif which is found towards the extracellular part of the M3 transmembrane domain. This motif forms the upper channel gate, which in the closed receptor contains all four Ala643 residues of the tetramer in close proximity [36]. As these Ala643 residues are tightly packed by the surrounding residues, the introduction of a larger sidechain (valine) is expected to destabilize the closed gate. It follows that channel closure by deactivation is likely to be less favorable and slower, while the degree of desensitization would be reduced. We observed these effects not only for homomeric GluA2(Q), but also for heteromeric receptors containing just two GluA2(R) A643V subunits, and for all receptors containing the auxiliary protein TARP γ2. The increased potency of glutamate at homomeric GluA2(Q) variant receptors might result from the destabilized gate being more fully opened by subsaturating glutamate concentrations [37]. Interestingly, A643V does not produce the constitutive channel gating that occurs with threonine variants at the analogous site in other subunits [6, 38, 39].

### Perampanel

Having identified A643V as a GoF variant, we investigated perampanel with a view to its possible therapeutic use. We found the drug to be much less potent on GluA2 A643V than on the wild-type receptors. This reduction is likely due to the proximity of the perampanel binding site to position 643 – any rearrangement of the outer M3 helix to accommodate Val643 will displace the adjacent Phe644 which is the most indispensable of the amino acids that form the perampanel binding site [16]. This means that in our patient, the inhibition of GluA2 A643V-containing AMPARs by perampanel would be expected to be less than that of AMPARs containing only wild-type GluA1-4 subunits.

Although drugs that bind to alternative parts of the AMPAR could, in theory, display improved selectivity against GluA2 A643V, no candidate molecules are available in the clinic. Interestingly, however, one component of the medium chain triglyceride (MCT) ketogenic diet, an established therapy for drug-resistant epilepsy [26], may act, in part, through inhibition of AMPARs. Specifically, seizure control by decanoic acid, a constituent of the MCT ketogenic diet [40], has been shown to involve inhibition of AMPARs through binding at a site distinct from that of perampanel [41]. Indeed, when used in combination, perampanel and decanoic acid act synergistically to inhibit AMPARs and provide enhanced suppression of seizure-like activity recorded *in vitro* from rodent and human brain tissue [42]. The current treatment plan for our patient – a modified Atkins diet (with MCT oil supplementation) alongside perampanel – would be expected to inhibit AMPARs through actions at two different sites, and this may contribute to its success despite the limited relative potency of perampanel against A643V variant receptors. However, it should be noted that in our patient, MCT oil supplementation of the modified Atkins diet was not initiated until 2 months after the introduction of perampanel. Another fatty acid, *trans*-4-butylcyclohexanxe carboxylic acid (4-BCCA), a cyclic derivative of octanoic acid, has also been shown to be effective against epileptiform activity *in vitro* [43] and bind to AMPARs at a site different from that of perampanel [44]. In future studies it will be of interest to assess the efficacy of a range of organic molecules including decanoic acid and 4-BCCA (alone or in combination with perampanel) as blockers of the A643V variant.

Genetic testing to identify monogenic epilepsies can potentially reveal appropriate therapeutic targets. Our study demonstrates that the potential benefits of such precision medicine can justify the time and effort involved. In our case, although previous *GRIA2* variants were identified as LoF [7], coupling the results of the extended genetic screen with detailed functional study highlighted the NAM perampanel as a logical candidate for therapy. Adding perampanel to our patient’s ongoing regimen was successful in reducing seizure number and has allowed weaning from levetiracetam (complete) and phenobarbital (ongoing). Notably, the gastrointestinal symptoms and failure to thrive also resolved. Whether this could reflect perampanel-inhibition of overactive variant GluA2-containing receptors, either centrally or in the peripheral/enteric nervous system, is unclear.

Commonly reported side-effects associated with perampanel use include impairments such as dizziness and sedation, as well as behavioral side effects such as negative mood alteration and aggression [45, 46]. Remarkably, our patient displayed quite the opposite behavior; he has shown increased alertness with improving fine motor control while his reported mood is significantly improved. A parsimonious explanation for this would be that, when typically prescribed, perampanel produces global underactivity of AMPARs while in a patient with AMPAR GoF it may instead promote a normalization of AMPAR-mediated excitation. Whatever the explanation, in the case of *GRIA2* GoF, perampanel not only reduces seizures but appears to facilitate developmental gain. This raises the possibility that other patients with *GRIA* GoF variants might benefit from perampanel treatment, even in the absence of epilepsy.

## Materials and Methods

### Clinical details

The study was deemed to be ‘not regulated’ by the Institutional Review Board (IRB) of the University of Michigan; publishing the clinical findings from a single individual does not fit the definition of human subjects research requiring IRB approval (per 45 CFR 46, 21 CFR 56 and University of Michigan policy). Written informed consent for genetic testing and publication was obtained from the patient’s parents. Trio genetic analysis was performed by GeneDX (Gaithersburg, MD) using the EpiXpanded Panel (https://www.genedx.com/tests/detail/epixpanded-panel-835). Treatment outcomes were obtained from neurological assessment, interviews with the patient’s parents, and evaluations by the patient’s occupational therapist.

### Heterologous expression

We expressed recombinant human AMPAR subunits and TARP γ2 (kind gifts from Dimitri Kullmann, UCL Queen Square Institute of Neurology, London, and Michael Maher, Janssen Research & Development L.L.C., San Diego, respectively), and EGFP, in HEK293 cells. These were maintained under standard protocols, as described previously [37]. GluA2 subunit cDNA was of the flip splice form and was R/G unedited. The GluA2 A643V point mutation was produced using standard PCR protocols. AMPAR/TARP combinations were transfected at a cDNA ratio of 1:1. For GluA1/2 receptors, the subunits were expressed at a ratio of 1:2. Transient transfection was performed using Lipofectamine 2000 (Life Technologies), and electrophysiological recordings were performed 18-48 h later.

### Electrophysiology

Recordings were made with an external solution containing 145 mM NaCl, 2.5 mM KCl, 1 mM CaCl_2_, 1 mM MgCl_2_, and 10 mM HEPES, pH 7.3. Patch-clamp electrodes were pulled from borosilicate glass (1.5 mm o.d., 0.86 mm i.d.; Harvard Apparatus) and fire polished to a final resistance of 8-12 MΩ. The internal solution contained 145 mM CsCl, 2.5 mM NaCl, 1 mM Cs-EGTA, 4 mM MgATP, and 10 mM HEPES (pH 7.3 with CsOH) and was supplemented with 100 μM spermine tetrahydrochloride or 100 μM NASPM for recordings from GluA2(Q) or GluA1/2, respectively. Recordings were made from outside-out patches at 22-25 °C using an Axopatch 200B amplifier (Molecular Devices). Currents were recorded at −60 mV for GluA2(Q) or +60 mV for GluA1/2, low-pass filtered at 10 kHz, and digitized at 20 kHz using an NI USB-6341 (National Instruments) interface with Strathclyde Electrophysiology Software WINWCP (John Dempster, University of Strathclyde, Glasgow UK).

### Rapid agonist application to excised patches

Rapid agonist application was achieved by switching between continuously flowing solutions. Solution exchange was achieved by moving an application tool made from theta glass (Hilgenberg) or triple barreled glass (Vitrocom) mounted on a piezoelectric translator (Physik Instrumente). The 10-90% exchange times, assessed by jumping open electrodes into a diluted solution and observing junction potential changes, were between 150 and 300 μs.

Experiments to assess dose response relationships for glutamate were performed using triple barreled glass. For experiments with cyclothiazide, each barrel contained a different glutamate concentration. In experiments to assess the glutamate *EC*_50_ of responses to fast jumps, the central barrel and left-hand barrel contained control and 10 mM glutamate solutions throughout, while the concentration in the right-hand barrel was varied. By alternately jumping into the streams from the left and right barrels, any effect of rundown was negated through normalization of the test response to the 10 mM glutamate response. Potency of inhibition by perampanel was assessed by comparing initial peak responses produced by 500 ms fast jumps into 10 mM glutamate in the absence or continuous presence of different concentrations of perampanel.

### Data analysis

Records were analyzed using Igor Pro 6.35 (Wavemetrics) with Neuromatic 2.8 [47]. Entry into desensitization (500 ms application of 10 mM glutamate) and current deactivation (1-2 ms application of 10 mM glutamate) were fitted with the sum of two exponentials and the weighted time constants (T_w, des_ and T_w, deact_) calculated, according to

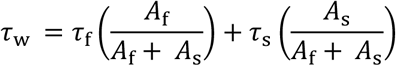

where *A*_*f*_ and *τ*_*f*_ are the amplitude and time constant of the fast component and *A*_*s*_ and *τ*_*s*_ are the amplitude and time constant of the slow component.

Non-stationary fluctuation analysis was performed on the decaying phase of currents evoked by 500 ms applications of 10 mM glutamate, as previously described [48]. The variance for each successive pair of current responses was calculated and the single-channel current (*i*) and total number of channels (*N*) were then determined by plotting the ensemble variance (σ^2^) against mean current (*Ï*) and fitting with a parabolic function:

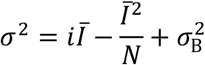

where σ_Β_^2^ is the background variance. The weighted mean single-channel conductance was calculated from the single-channel current and the holding potential.

*EC*_50_ and *IC*_50_ values were derived by fitting the Hill equation:

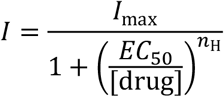

where *I*_max_ is the peak of the fit, *EC*_50_ is the concentration producing the half-maximal response, *n*_H_ is the Hill coefficient and [drug] refers to the concentration of either glutamate or perampanel. Before pooling the data, the *I*_max_ value derived from the fit for each patch was used to rescale the individual datasets. The rescaled values were then averaged and refit for display purposes only.

### Data presentation and statistical analysis

Statistical analysis was performed using R (version 4.1.1, the R Foundation for Statistical Computing, https://www.r-project.org/) and R Studio (version 1.4.1717, RStudio). Summary data are presented in the text as mean ± standard error (s.e.m.) from n patches. The data for the different measures from wild-type and variant receptors are also presented in **Table 1** and **Table 2**, together with unpaired mean differences and their 95% confidence intervals. Bias corrected and accelerated (BCa) confidence intervals were calculated from 5000 bootstrap resamples using the dabestr package in R [49]. Normality was not tested statistically but gauged from density histograms and/or quantile-quantile plots. All p-values were calculated using a non-parametric two-sided approximate permutation t-test, with 10000 bootstrap replicates using the Coin package in R [50]. No statistical test was used to predetermine sample sizes; these were based on standards of the field.

## Data Availability

All data produced are available online at figshare.

https://doi.org/10.5522/04/19279406

## Acknowledgements

This work was supported by a Medical Research Council Programme Grant to Mark Farrant and Stuart G. Cull-Candy [Grant MR/T002506/1].

We thank the patient’s family for their generous collaboration and the CureGRIN Foundation (https://curegrin.org) for facilitating this interaction. We are grateful for the assistance of Casey Bross. We thank Dimitri Kullmann, Michael Maher and Henry Holden for helpful materials and/or discussion.

## Supplementary Material

**Figure S1:**
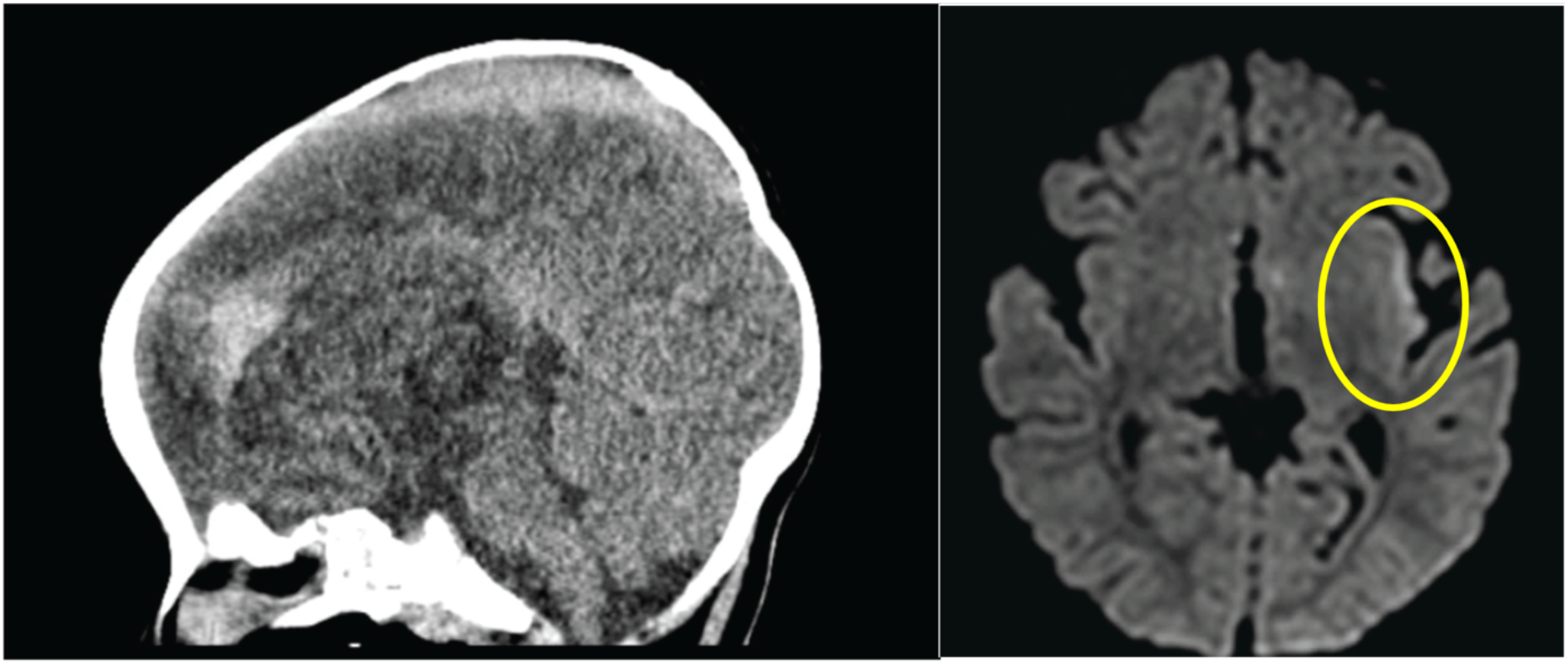
Head imaging at 9 months of age upon presentation for seizures. Sagittal CT image (left) showed extra-axial hyperdensity along the anterior falx, concerning for a subdural hematoma. Diffusion weighted axial MRI imaging (right) was notable for diffusion restriction along the left insular cortex, likely due to ongoing seizures.

